# Glucose Dispersion Is Associated With Contrast Sensitivity Beyond Average Glycemia

**DOI:** 10.64898/2026.09.08.26362509

**Authors:** Ren Zhang

## Abstract

**Background:** HbA1c and mean glucose summarize average glycemia but do not capture glucose dispersion. Whether free-living glucose dispersion is associated with visual function beyond average glycemia remains unclear.

**Aim:** To determine whether glucose dispersion is associated with mesopic and photopic contrast sensitivity independently of HbA1c and contemporaneous CGM mean glucose.

**Methods:** We analyzed 2,218 AI-READI participants with continuous glucose monitoring and visual-function data. Glucose dispersion was represented primarily by mean absolute deviation from mean glucose (AAD), and associations were assessed using multivariable linear regression.

**Results:** Higher AAD was associated with lower mesopic and photopic contrast sensitivity after adjustment for age and HbA1c (β = −0.188 and −0.165; P = 1.28 × 10⁻¹¹ and 8.55 × 10⁻⁹) and after adjustment for age and CGM mean glucose (β = −0.221 and −0.195; P = 4.84 × 10⁻¹³ and 5.66 × 10⁻¹⁰). Associations persisted with additional adjustment and sensitivity analyses; glucose SD yielded concordant results.

**Conclusion:** Glucose dispersion is associated with contrast sensitivity beyond average glycemia, suggesting that glycemic exposure may relate to visual function through its average level and dispersion.

## 1. Introduction

Diabetes is a systemic metabolic disease whose long-term burden is driven in large part by chronic vascular and neural complications, including diabetic eye disease [1, 2]. Although diabetic retinopathy is clinically defined primarily by retinal vascular abnormalities, visual dysfunction may extend beyond structural changes and high-contrast visual acuity. Contrast sensitivity, which reflects the ability to distinguish luminance differences between an object and its background, provides a complementary measure of visual function. Reduced contrast sensitivity has been reported in people with diabetes, including in individuals without clinically apparent retinopathy, and may be particularly evident under mesopic conditions [2–4]. These observations suggest that functional visual measures may capture aspects of diabetes-related ocular dysfunction not fully reflected by conventional clinical endpoints.

Glycemic exposure is a major determinant of diabetes-related microvascular complications [1], but it is multidimensional. HbA1c remains the standard measure of longer-term average glycemia, whereas mean glucose summarizes average glycemic exposure over a defined observation period [5]. Neither measure, however, describes how widely glucose values fluctuate around that average. Continuous glucose monitoring (CGM) provides dense glucose measurements under free-living conditions and permits characterization of mean glucose, time within and outside target ranges, and glycemic variability [6]. Moreover, similar HbA1c values can correspond to substantially different mean glucose concentrations and glucose profiles [7]. Thus, average glycemia and glucose dispersion represent related but distinct features of glycemic exposure. Prior studies have linked poorer glycemic status with reduced contrast sensitivity and have examined CGM-derived measures of glycemic variability or time in range in relation to diabetic retinopathy, retinal structure, and retinal functional measures [8–10]. Direct evidence relating free-living CGM-derived glucose dynamics to contrast sensitivity, however, remains limited [8]. In particular, it remains unclear whether glucose dispersion is associated with mesopic and photopic contrast sensitivity after accounting for both longer-term glycemia and contemporaneous mean glucose in a large population spanning a broad range of metabolic states.

We therefore examined the association between CGM-derived glucose dispersion and mesopic and photopic contrast sensitivity in the AI-READI cohort, which combines standardized visual-function testing with free-living CGM across diverse metabolic states [11, 12]. Glucose dispersion was characterized primarily using mean absolute deviation from mean glucose (AAD), with glucose SD retained as a conventional variability benchmark. We tested whether greater glucose dispersion was associated with poorer contrast sensitivity after adjustment for age and HbA1c and, separately, for age and contemporaneous CGM mean glucose, and examined these associations across metabolic groups.

## 2. Materials and methods

### 2.1 Study design, data source, and analytic cohort

This study was a cross-sectional secondary analysis of the AI-READI (Artificial Intelligence– Ready and Exploratory Atlas for Diabetes Insights) Flagship Dataset of Type 2 Diabetes, version 3.0.0. AI-READI is a prospective, multicenter study conducted at three U.S. sites and designed to collect deeply phenotyped, multimodal data from adults with and without type 2 diabetes across a range of metabolic states. The current data release includes 2,280 participants and integrates laboratory measurements, vision assessments, continuous glucose monitoring (CGM), imaging, wearable-sensor data, and other phenotypic domains.

The analytic cohort was constructed by integrating participant-level CGM data with validated eye and metabolic data. Of the 2,280 participants in the release, 2,245 had raw CGM data and 2,240 contributed at least one quality-control–valid CGM day. After linkage with eye and metabolic data, 2,218 participants constituted the descriptive CGM–eye cohort. Mesopic and photopic contrast-sensitivity measurements were available for 2,210 and 2,214 participants, respectively. HbA1c was available for 2,155 participants; therefore, the HbA1c-adjusted analyses included 2,147 participants for mesopic contrast sensitivity and 2,151 for photopic contrast sensitivity. Model-specific complete-case samples were used for analyses not requiring HbA1c. The present study was approved by the Institutional Review Board of Wayne State University.

### 2.2 Visual outcomes, metabolic characterization, and CGM measures

Mesopic and photopic contrast sensitivity were assessed monocularly using the Mars Letter Contrast Sensitivity Test and expressed as log contrast sensitivity (logCS). For each lighting condition, the primary participant-level outcome was the mean of available right- and left-eye measurements; when only one eye was available, that measurement was retained. Analyses restricted to participants with measurements from both eyes were performed as sensitivity analyses.

Age, HbA1c, and metabolic classification were obtained from the validated AI-READI data. Participants were classified according to the four AI-READI metabolic groups: healthy, prediabetes/lifestyle, non-insulin medication, and insulin-dependent. These variables were used for cohort characterization, covariate adjustment, and subgroup analyses as appropriate. CGM data were obtained from Dexcom G6 recordings. Glucose values were standardized to mg/dL, timestamped, deduplicated, and subjected to day-level quality control. A QC-valid day required at least 1,000 minutes of valid CGM data. Participant-level glycemic measures were calculated from QC-valid days and included mean glucose, time in range (70–180 mg/dL), time above range (>180 mg/dL; TAR180), glucose SD, and mean absolute deviation from mean glucose (AAD). For glucose observations *G_i_*and participant-level mean glucose *G̅*, AAD was defined as

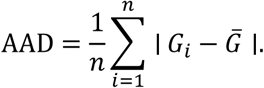

### 2.3 Statistical analysis

Continuous variables were summarized as median [Q1, Q3] and categorical variables as n (%). Associations of AAD with mesopic and photopic contrast sensitivity were examined separately using linear regression. Within each model-specific complete-case sample, AAD, the contrast-sensitivity outcome, and continuous covariates were standardized to mean 0 and SD 1. Thus, β represents the adjusted SD difference in contrast sensitivity associated with a 1-SD higher AAD. Results are reported as standardized regression coefficients (β), 95% confidence intervals, P values, and incremental R² (ΔR²), defined as the increase in R² after adding AAD to the corresponding covariate-only model. Primary models adjusted for age and HbA1c or for age and CGM mean glucose, representing longer-term and contemporaneous average glycemia, respectively. Additional models adjusted for age, HbA1c, and TAR180; age, HbA1c, and metabolic group; or age, HbA1c, CGM mean glucose, and metabolic group. Model-specific complete cases were used.

Associations were also examined within each metabolic group after adjustment for age and CGM mean glucose, with effect modification assessed using an AAD × metabolic-group interaction term. For visualization, adjusted contrast-sensitivity means were estimated across AAD deciles, whereas statistical inference treated AAD as a continuous variable. AAD was selected as the primary glucose-dispersion measure following an exploratory comparison of candidate measures that considered association strength across both contrast-sensitivity outcomes, cross-validated performance, and measurement reliability; glucose SD was retained as a conventional benchmark. Sensitivity analyses examined alternative visual-outcome definitions and model specifications. All tests were two-sided, and P<0.05 was considered statistically significant. Analyses were performed using Python 3.12.3, with ordinary least-squares models fitted using statsmodels 0.15.0; figures were generated using Matplotlib.

## 3. Results

### 3.1 Analytic cohort and glycemic characteristics

To characterize the analytic cohort and its glycemic profile across metabolic states, we first examined cohort composition and key measures of average glycemia and glucose dispersion. Of 2,280 participants in the AI-READI release, 2,245 had raw CGM data and 2,240 contributed at least one QC-valid CGM day. After integration with eye and metabolic data, 2,218 participants comprised the descriptive analytic cohort; mesopic and photopic contrast-sensitivity measurements were available for 2,210 and 2,214 participants, respectively. The median age was 61 [52, 69] years. The cohort included 751 (33.9%) healthy participants, 550 (24.8%) in the prediabetes/lifestyle group, 672 (30.3%) receiving non-insulin medication, and 245 (11.0%) in the insulin-dependent group.

Median HbA1c was 5.80 [5.50, 6.30]%, and median CGM mean glucose was 124.9 [113.2, 145.2] mg/dL. Both measures increased progressively across the four metabolic groups. Glucose dispersion was characterized primarily using mean absolute deviation from mean glucose (AAD), with glucose SD retained as a conventional variability measure. Median AAD was 17.3 [13.8, 24.6] mg/dL and median glucose SD was 22.8 [18.5, 31.9] mg/dL; participants contributed a median of 10 [9, 10] QC-valid CGM days. Median mesopic and photopic contrast sensitivities were 1.30 [1.18, 1.42] and 1.60 [1.48, 1.68] logCS, respectively. As an illustrative example, two participants with nearly identical mean glucose (125.1 versus 124.8 mg/dL) had markedly different AAD values (15.3 versus 34.1 mg/dL), demonstrating substantial differences in glucose dispersion despite similar average glycemia.

### 3.2 Glucose dispersion is associated with contrast sensitivity beyond average glycemia

We next asked whether glucose dispersion was associated with contrast sensitivity beyond differences in average glycemia. This distinction was central because greater glucose variability can accompany higher overall glycemic exposure; an association with contrast sensitivity could therefore reflect average hyperglycemia rather than dispersion itself. We first adjusted for age and HbA1c, a measure of longer-term average glycemia. Higher AAD remained strongly associated with lower mesopic contrast sensitivity (β = −0.188, 95% CI −0.242 to −0.134; P = 1.28 × 10⁻¹¹) and lower photopic contrast sensitivity (β = −0.165, 95% CI −0.221 to −0.109; P = 8.55 × 10⁻⁹) (Figure 2A,B; Table 2). Adding AAD increased model R² by 0.0174 for mesopic and 0.0134 for photopic contrast sensitivity, corresponding to 1.74 and 1.34 percentage points of additional explained variance beyond age and HbA1c.

**Figure 1.**
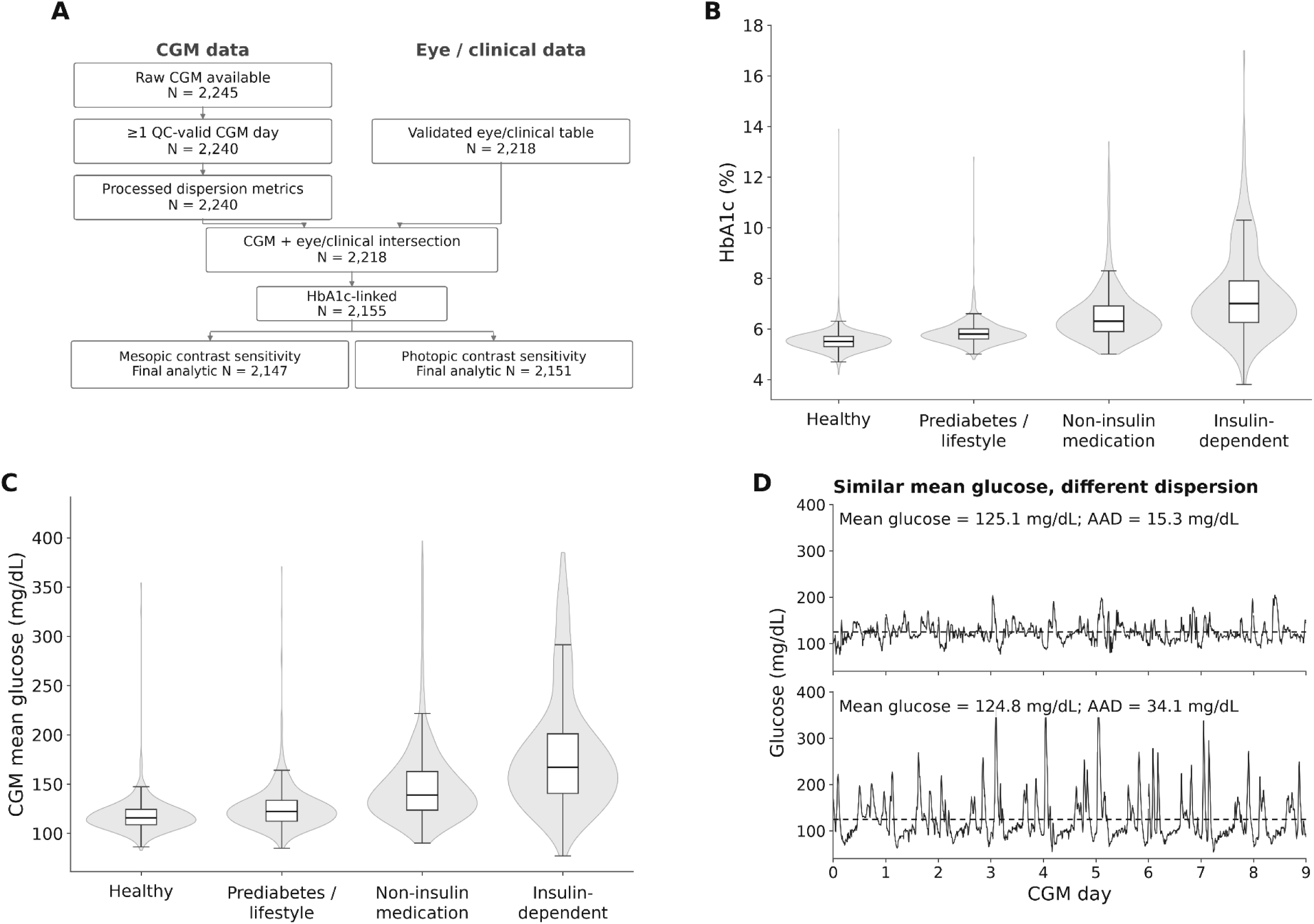
AI-READI analytic cohort and glycemic characteristics. (A) Analytic flow diagram showing integration of CGM and eye/clinical data and derivation of the mesopic and photopic contrast-sensitivity samples. Distributions of (B) HbA1c and (C) CGM mean glucose across the four metabolic groups. (D) Outcome-blind illustrative CGM profiles from two participants with nearly identical mean glucose but markedly different mean absolute deviation from mean glucose (AAD), illustrating differences in glucose dispersion despite similar average glycemia.

**Figure 2.**
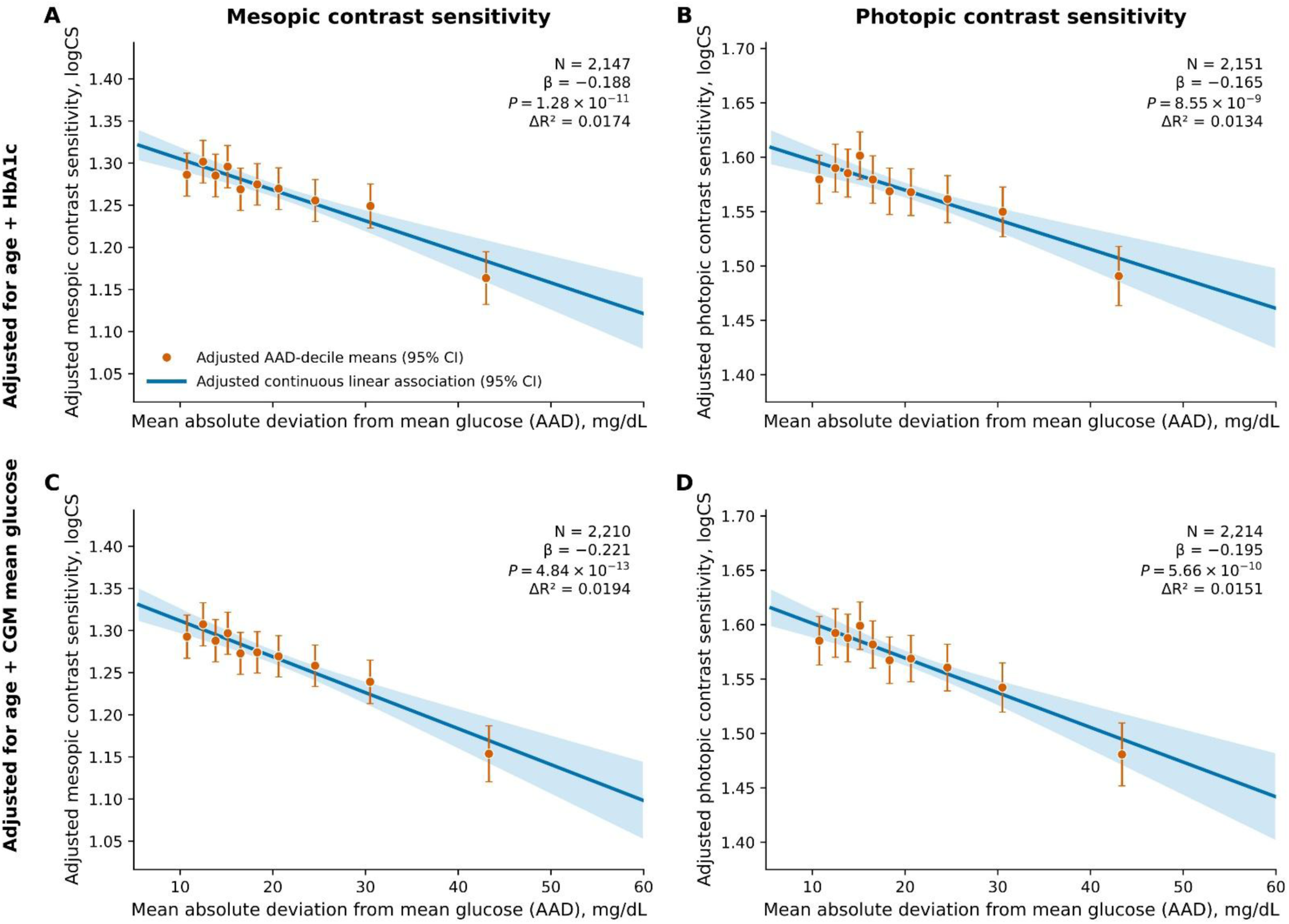
Glucose dispersion and contrast sensitivity after adjustment for HbA1c or mean glucose. Associations of mean absolute deviation from mean glucose (AAD) with mesopic (A, C) and photopic (B, D) contrast sensitivity were examined after adjustment for age and HbA1c (A, B) or age and CGM mean glucose (C, D). Orange points show adjusted means within AAD deciles with 95% CIs; blue lines and bands show continuous adjusted linear associations with 95% CIs. Standardized β, *P*, and ΔR² are shown for each model; ΔR² represents the additional variance explained by AAD beyond the corresponding base model.

**Figure 3.**
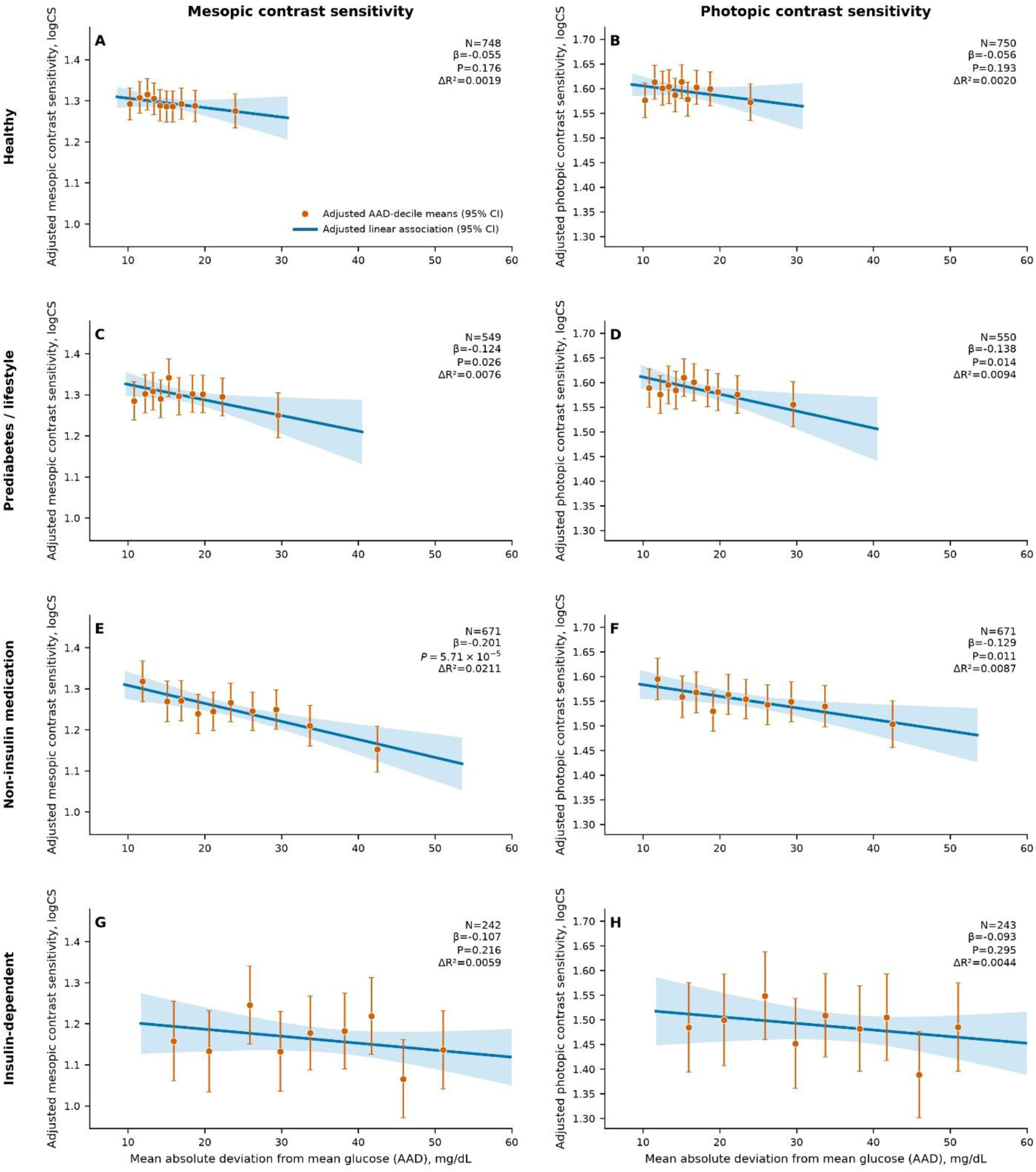
Glucose dispersion and contrast sensitivity across metabolic groups. Associations of mean absolute deviation from mean glucose (AAD) with mesopic and photopic contrast sensitivity are shown within healthy, prediabetes/lifestyle, non-insulin medication, and insulin-dependent groups after adjustment for age and CGM mean glucose. Orange points indicate adjusted means within AAD deciles for each metabolic group with 95% confidence intervals; blue lines and bands show continuous adjusted linear associations with 95% confidence intervals. Standardized β, *P*, and ΔR² are shown for each subgroup. Global AAD × metabolic-group interaction tests showed no significant effect modification for mesopic (*P* = 0.243) or photopic (*P* = 0.302) contrast sensitivity.

**Table 1.** Analytic cohort characteristics.

| **Characteristic** | **N** | **Median [Q1, Q3] / n (%)** |
| --- | --- | --- |
| **Cohort** |  |  |
| Participants | 2,218 |  |
| **Age** |  |  |
| Age, years | 2,218 | 61 [52, 69] |
| **Metabolic group** |  |  |
| Healthy |  | 751 (33.9) |
| Prediabetes / lifestyle |  | 550 (24.8) |
| Non-insulin medication |  | 672 (30.3) |
| Insulin-dependent |  | 245 (11.0) |
| **Average glycemia** |  |  |
| HbA1c (%) | 2,155 | 5.80 [5.50, 6.30] |
| CGM mean glucose (mg/dL) | 2,218 | 124.9 [113.2, 145.2] |
| **Glucose dispersion** |  |  |
| Mean absolute deviation from mean glucose (AAD), mg/dL | 2,218 | 17.3 [13.8, 24.6] |
| Glucose SD (mg/dL) | 2,218 | 22.8 [18.5, 31.9] |
| **Glycemic range / burden** |  |  |
| Time in range, 70–180 mg/dL (%) | 2,218 | 96.3 [86.4, 98.9] |
| Time above range, >180 mg/dL (%) | 2,218 | 2.5 [0.5, 12.8] |
| **CGM coverage** |  |  |
| QC-valid CGM days | 2,218 | 10 [9, 10] |
| **Visual outcomes** |  |  |
| Mesopic contrast sensitivity (logCS) | 2,210 | 1.30 [1.18, 1.42] |
| Photopic contrast sensitivity (logCS) | 2,214 | 1.60 [1.48, 1.68] |

Values are median [Q1, Q3] unless otherwise indicated. The descriptive cohort is the validated CGM–eye/clinical intersection (N=2,218). A QC-valid CGM day required ≥1,000 valid CGM minutes. Variable-specific N is shown for continuous measures; metabolic groups are n (%).

**Table 2.** Association of glucose dispersion with mesopic and photopic contrast sensitivity.

| Adjustment model | Mesopic contrast sensitivity |  |  |  | Photopic contrast sensitivity |  |  |  |
| --- | --- | --- | --- | --- | --- | --- | --- | --- |
| | N | Standardized $\beta$ (95% CI) | P | $\Delta R^2$ | N | Standardized $\beta$ (95% CI) | P | $\Delta R^2$ |
| **AAD models** |  |  |  |  |  |  |  |  |
| Age + HbA1c | 2,147 | -0.188 (-0.242, -0.134) | $1.28 \times 10^{-11}$ | 0.0174 | 2,151 | -0.165 (-0.221, -0.109) | $8.55 \times 10^{-9}$ | 0.0134 |
| Age + CGM mean glucose | 2,210 | -0.221 (-0.281, -0.162) | $4.84 \times 10^{-13}$ | 0.0194 | 2,214 | -0.195 (-0.257, -0.134) | $5.66 \times 10^{-10}$ | 0.0151 |
| Age + HbA1c + TAR180 | 2,147 | -0.215 (-0.283, -0.147) | $5.97 \times 10^{-10}$ | 0.0145 | 2,151 | -0.183 (-0.254, -0.113) | $3.38 \times 10^{-7}$ | 0.0105 |
| Age + HbA1c + metabolic group | 2,147 | -0.130 (-0.189, -0.071) | $1.48 \times 10^{-5}$ | 0.0070 | 2,151 | -0.104 (-0.165, -0.043) | $8.76 \times 10^{-4}$ | 0.0044 |
| Age + HbA1c + CGM mean glucose + metabolic group | 2,147 | -0.151 (-0.217, -0.084) | $8.81 \times 10^{-6}$ | 0.0074 | 2,151 | -0.122 (-0.191, -0.053) | $5.02 \times 10^{-4}$ | 0.0048 |
| **Conventional glucose SD benchmark** |  |  |  |  |  |  |  |  |
| Age + HbA1c | 2,147 | -0.177 (-0.230, -0.124) | $6.78 \times 10^{-11}$ | 0.0162 | 2,151 | -0.157 (-0.212, -0.102) | $2.30 \times 10^{-8}$ | 0.0126 |

AAD, mean absolute deviation from mean glucose. $\beta$ is the standardized regression coefficient for AAD; the benchmark row reports the standardized coefficient for glucose SD.
$\Delta R^2$ is the increase in model $R^2$ after adding AAD, or SD for the benchmark row, to the listed covariates.
TAR180 is the percentage of CGM time >180 mg/dL. Metabolic group is the validated four-level AI-READI variable.
The age + CGM mean-glucose model uses its natural larger complete-case sample; models containing HbA1c use the HbA1c-complete sample.

We then addressed average glycemia more directly using CGM mean glucose measured during the same monitoring period from which AAD was derived. After adjustment for age and CGM mean glucose, the inverse associations remained evident and were of similar or greater magnitude for mesopic (β = −0.221, 95% CI −0.281 to −0.162; P = 4.84 × 10⁻¹³) and photopic contrast sensitivity (β = −0.195, 95% CI −0.257 to −0.134; P = 5.66 × 10⁻¹⁰) (Figure 2C,D; Table 2). AAD contributed an additional 0.0194 and 0.0151 to R² for mesopic and photopic contrast sensitivity, respectively. Across both adjustment strategies, adjusted AAD-decile means followed the same inverse pattern as the continuous regression estimates, with greater glucose dispersion corresponding to poorer contrast sensitivity under both mesopic and photopic conditions. The persistence of the association after adjustment for both HbA1c and contemporaneous CGM mean glucose indicates that the visual-function relationship was not explained by average glycemia alone.

### 3.3 Associations across metabolic groups

We next examined whether the association between glucose dispersion and contrast sensitivity was present within individual metabolic groups after adjustment for age and CGM mean glucose. For mesopic contrast sensitivity, higher AAD was significantly associated with lower contrast sensitivity in the prediabetes/lifestyle group (β = −0.124, 95% CI −0.233 to −0.015; P = 0.026) and in the non-insulin medication group (β = −0.201, 95% CI −0.299 to −0.104; P = 5.71 × 10⁻⁵). Associations were also negative in the healthy and insulin-dependent groups but did not reach statistical significance.

A similar pattern was observed for photopic contrast sensitivity. Higher AAD was significantly associated with lower contrast sensitivity in the prediabetes/lifestyle group (β = −0.138, 95% CI −0.248 to −0.028; P = 0.014) and the non-insulin medication group (β = −0.129, 95% CI −0.229 to −0.030; P = 0.011), whereas associations in the healthy and insulin-dependent groups were not statistically significant. Formal interaction testing did not provide evidence that the association differed across metabolic groups for either mesopic (*P* = 0.243) or photopic (*P* = 0.302) contrast sensitivity. Thus, the strongest subgroup evidence was observed in the prediabetes/lifestyle and non-insulin medication groups, in which greater glucose dispersion was associated with poorer mesopic and photopic contrast sensitivity after accounting for age and contemporaneous mean glucose.

### 3.4 Associations after additional glycemic and metabolic adjustment

We further examined whether the association between glucose dispersion and contrast sensitivity persisted after accounting for other aspects of glycemic burden and metabolic status. After adjustment for age, HbA1c, and TAR180, higher AAD remained associated with lower mesopic contrast sensitivity (β = −0.215, 95% CI −0.283 to −0.147; P = 5.97 × 10⁻¹⁰; ΔR² = 0.0145) and lower photopic contrast sensitivity (β = −0.183, 95% CI −0.254 to −0.113; P = 3.38 × 10⁻⁷; ΔR² = 0.0105). Adjustment for metabolic group attenuated the associations, but both remained statistically significant. In models including age, HbA1c, and metabolic group, β was −0.130 for mesopic contrast sensitivity (P = 1.48 × 10⁻⁵; ΔR² = 0.0070) and −0.104 for photopic contrast sensitivity (P = 8.76 × 10⁻⁴; ΔR² = 0.0044). Associations also remained significant in the more comprehensive model including age, HbA1c, CGM mean glucose, and metabolic group (mesopic: β = −0.151, P = 8.81 × 10⁻⁶, ΔR² = 0.0074; photopic: β = −0.122, P = 5.02 × 10⁻⁴, ΔR² = 0.0048).

To determine whether the findings depended on the choice of AAD, glucose SD was examined as a conventional variability measure. After adjustment for age and HbA1c, higher glucose SD was similarly associated with lower mesopic (β = −0.177, P = 6.78 × 10⁻¹¹; ΔR² = 0.0162) and photopic contrast sensitivity (β = −0.157, P = 2.30 × 10⁻⁸; ΔR² = 0.0126). Together, these analyses support a robust association between glucose dispersion and contrast sensitivity that is not explained solely by hyperglycemic burden, metabolic group, or the specific dispersion metric used.

### 3.5 Sensitivity analyses

Sensitivity analyses supported the robustness of the primary findings. Restricting the analysis to participants with measurements available from both eyes produced estimates similar to the primary models for mesopic (N = 2,132; β = −0.184, 95% CI −0.239 to −0.129; P = 5.61 × 10⁻¹¹; ΔR² = 0.0164) and photopic contrast sensitivity (N = 2,138; β = −0.163, 95% CI −0.220 to −0.106; P = 2.00 × 10⁻⁸; ΔR² = 0.0128). Results were also preserved using HC3 robust standard errors (mesopic: β = −0.188, P = 3.01 × 10⁻¹⁰; photopic: β = −0.165, P = 7.60 × 10⁻⁸) and after exclusion of influential observations based on Cook’s distance (mesopic: β = −0.181, P = 3.19 × 10⁻¹¹; photopic: β = −0.172, P = 5.67 × 10⁻¹⁰).

Requiring at least nine QC-valid CGM days likewise yielded similar associations (mesopic: N = 1,998; β = −0.179, P = 2.65 × 10⁻¹⁰; photopic: N = 1,999; β = −0.154, P = 1.46 × 10⁻⁷). Natural-spline analyses confirmed significant overall associations for both mesopic (P = 1.29 × 10⁻¹⁰) and photopic contrast sensitivity (P = 8.70 × 10⁻⁹). There was no significant evidence of nonlinearity for mesopic contrast sensitivity (P = 0.097), whereas the photopic association showed evidence of departure from linearity (P = 0.017). Overall, the inverse association between glucose dispersion and contrast sensitivity was robust to alternative outcome definitions, inference methods, influential observations, and stricter CGM coverage requirements.

## 4. Discussion

In this large, metabolically diverse cohort, greater free-living glucose dispersion was associated with poorer mesopic and photopic contrast sensitivity. Importantly, these associations persisted after accounting for two complementary measures of average glycemia: HbA1c, reflecting longer-term glycemic exposure, and CGM mean glucose, measured during the same observation period from which glucose dispersion was derived. The associations also remained after additional adjustment for hyperglycemic burden and metabolic group and were reproduced using conventional glucose SD. Taken together, these findings suggest that the relationship between glycemia and visual function is not fully captured by average glucose exposure alone. Rather, the distribution of glucose values around that average may represent an additional dimension relevant to visual function.

Glucose dispersion reflects the spread of an individual’s observed glucose values around their average level. Individuals with similar HbA1c or mean glucose may therefore experience substantially different distributions of glucose exposure. Such differences could reflect patterns of glycemic exposure that are not represented by average glycemia alone. In human studies, acute glucose fluctuations have been associated with increased oxidative stress, while oxidative stress, inflammation, mitochondrial dysfunction, endothelial injury, and impaired neurovascular coupling are well-established components of diabetic retinal disease [13–16]. The retina is particularly dependent on tightly coordinated neural, glial, and vascular responses to changing metabolic demand, and impairment of this neurovascular unit can occur early in diabetes [14, 15]. These pathways provide biologically plausible explanations for an association between glucose dispersion and contrast sensitivity, although the present cross-sectional study cannot determine whether glucose fluctuations themselves contribute causally to visual dysfunction.

Notably, the strongest subgroup evidence was observed in the prediabetes/lifestyle and non-insulin medication groups, whereas estimates in the healthy and insulin-dependent groups were negative but less precise. Retinal functional abnormalities have previously been reported in prediabetes and early diabetes [17], making the presence of an association in intermediate metabolic groups of particular interest. However, these metabolic groups should not be interpreted as sequential disease stages, and the subgroup findings require confirmation.

Several features strengthen the study. AI-READI provided a large cohort spanning a broad range of metabolic states, standardized assessment of both mesopic and photopic contrast sensitivity, and free-living CGM measurements. The availability of both HbA1c and contemporaneous CGM mean glucose allowed glucose dispersion to be evaluated against complementary measures of average glycemia. Results were consistent across multiple adjustment strategies and remained robust to alternative visual-outcome definitions, robust inference, exclusion of influential observations, and stricter CGM coverage requirements. The similar findings obtained with glucose SD further support interpretation of the results as a glucose-dispersion phenomenon rather than one dependent on a particular summary measure.

Several limitations should also be considered. The cross-sectional design precludes inference about temporal direction or causality. CGM characterized glucose exposure over a relatively short period and may not fully represent an individual’s longer-term pattern of glucose dispersion, although most participants contributed approximately 9–10 valid days and results were preserved under stricter coverage requirements. Residual confounding cannot be excluded despite adjustment for major measures of glycemic exposure and metabolic status. Contrast sensitivity is a global functional measure and does not localize dysfunction to a particular retinal cell type, vascular compartment, or visual pathway. In addition, AAD was selected through exploratory comparison of candidate dispersion measures and should not be interpreted as an established optimal metric; the concordant SD results indicate that the broader physiological construct of glucose dispersion is more important than the specific measure used here. Finally, subgroup estimates differed in precision because of unequal group sizes and heterogeneity of metabolic status and treatment.

## Conclusion

Greater glucose dispersion was associated with poorer mesopic and photopic contrast sensitivity beyond both HbA1c and contemporaneous mean glucose. These findings suggest that glycemic exposure may be relevant to visual function not only through its average level, but also through the dispersion of glucose over time. Prospective studies are needed to determine whether glucose dispersion contributes to the development or progression of visual dysfunction.

## Statements and Declarations

### Ethical approval

The present study was approved by the Institutional Review Board of Wayne State University.

### Consent to Participate

Not applicable.

### Author Contributions

R.Z. conceptualized, designed the study, analyzed the data, and wrote the paper.

### Ethical Responsibilities of Authors

The author has read, understood, and complied, as applicable, with the statement on Ethical Responsibilities of Authors in the Instructions for Authors.

### Data Availability Statement

The AI-READI Flagship Dataset of Type 2 Diabetes is publicly available through the AI-READI data portal at aireadi.org, with dataset access provided through FAIRhub. The present study used version 3.0.0 of the dataset.

